# Optimization of Super disintegrants in Clonazepam Orally Fast Disintegrating Tablets: Impact on Dissolution, Drug Release and Stability

**DOI:** 10.1101/2025.03.18.25324237

**Authors:** Samira Karim, Zubair Khalid Labu, Shakil Khan, Sidratul Muntaha Ritu, Sraboni Mim, Md. Abdullah Sarker, Md. Tarekur Rahman

## Abstract

The present study systematically investigated the role of sodium starch glycolate, kollidon CL, and ludiflash in nine (9) formulations (F-1 to F-9) of clonazepam orally fast disintegrating tablets (OFDT) developed via direct compression. Advanced preformulating assessments ensured optimal flow properties and compressibility, facilitating robust tablet manufacturing. *In vitro* dissolution studies revealed that formulations incorporating different superdisintegrants (F-3, F-6, and F-9) exhibited superior drug release profiles with faster drug release, attributed to the concentration and characteristics of the excipients, with F-5 demonstrating the highest dissolution efficiency (94%). Drug release kinetics followed first-order models with anomalous (non-Fickian) diffusion mechanisms, highlighting the interplay between formulation composition and drug release behavior. Similarity factor (f_2_) analysis confirmed moderate alignment with the marketed product, identifying key areas for further optimization. Stability testing, conducted under ICH guidelines, demonstrated the long-term integrity of all formulations. This study bridges a critical research gap by providing an in-depth analysis of superdisintegrants selection, dissolution behavior, and stability, offering a strategic approach for optimizing OFDT formulations for improved therapeutic efficacy and patient compliance.

**Author Summary:** Optimization of superdisintegrants in clonazepam orally fast disintegrating tablets : Impact on dissolution, drug release, and stability is based on the pharmaceutical formulation and development of OFDT of clonazepam, a benzodiazepine used for treating epilepsy and panic disorders. The research aims to enhance the disintegration efficiency, dissolution rate, and bioavailability of the drug by incorporating three superdisintegrants: sodium starch glycolate, kollidon CL, and ludiflash through the direct compression method, a widely used pharmaceutical manufacturing process. The study involves preformulation analysis, physicochemical evaluation, *in vitro* dissolution studies, and stability testing under ICH guidelines, ultimately identifying F-5 as the most effective formulation with 94% dissolution efficiency. The findings contribute to the optimization of excipient selection in fast-dissolving formulations to improve the therapeutic performance and patient compliance of clonazepam.

## 1. Introduction

The primary objective of any drug delivery system is to transport an appropriate dose of the drug to the target site within the body, ensuring the desired concentration is reached swiftly and sustained throughout the treatment period. An effective system must release the drug at a rate that corresponds to the body’s requirements during the therapy [1].

In contemporary pharmaceutical technology, various drug delivery systems are available. Among these, oral administration is the most favored method for delivering drugs systematically [2]. This preference is attributed to its ease of ingestion, accurate dosing, convenience for self-administration, avoidance of pain, versatility, minimal discomfort, and, most importantly, enhanced patient compliance [3, 4]. Tablets and capsules are the most widely used oral dosage forms. However, a major limitation associated with these forms is dysphagia, or difficulty swallowing, which affects approximately 35% of the population [5].

To overcome the challenge of swallowing difficulties, pharmaceutical experts have introduced fast-dissolving drug delivery systems, such as mouth-dissolving tablets. These tablets disintegrate and dissolve in saliva within seconds, eliminating the need for water or chewing. Typically, a mouth-dissolving tablet dissolves within 15 seconds to 3 minutes. Recent innovations in novel drug delivery systems (NDDS) aim to improve drug safety and reduce toxicity by developing user-friendly dosage forms that enhance patient compliance. One such advancement is the creation of fast-dissolving tablets [6].

Fast-dissolving drug delivery systems, first developed in the late 1970s, provide an effective alternative to conventional dosage forms, especially for pediatric and geriatric patients who struggle with swallowing traditional tablets and capsules. These tablets dissolve in saliva, often within 60 seconds, making them highly convenient for individuals with dysphagia. Fast-dissolving/disintegrating tablets (FDDTs) are also referred to as fast-melting, quick-dissolving, rapid-dispersing, and rapid-melt tablets. The U.S. Food and Drug Administration (FDA) categorize all FDDTs as orally disintegrating tablets. Additionally, the European Pharmacopoeia defines “Orodispersible Tablets” as those that rapidly disperse in the oral cavity before being swallowed.

The key benefit of FDDTs lies in their ability to merge the advantages of both liquid and conventional tablet forms. This unique formulation allows for the convenience of a solid dosage while ensuring the ease of swallowing associated with liquid medications. Additionally, FDDTs offer more accurate dosing compared to liquid oral formulations [7]. Epileptic seizures are characterized by sudden and repetitive sensory disturbances, which can lead to loss of consciousness or convulsions due to abnormal electrical activity in the brain. Notably, around 90% of epilepsy cases are reported in developing nations [8].

Clonazepam, a high-potency benzodiazepine classified as a class II agent, is FDA-approved for the treatment of status epilepticus and panic disorders [9]. Its chemical name, according to IUPAC, is 5-(2-chlorophenyl)-1,3-dihydro-7-nitro-2H-1,4-benzodiazepin-2-one. Clonazepam appears as a light-yellow crystalline powder with a molecular weight of 315.72 g/mol.

Following oral administration, Clonazepam is rapidly and fully absorbed, with an absolute bioavailability of approximately 90%. Peak plasma concentrations are typically achieved within 1 to 4 hours. Around 85% of the drug binds to plasma proteins, and it undergoes extensive metabolism, with less than 2% of the unchanged drug excreted in the urine. The elimination half-life ranges between 30 to 40 hours, and its pharmacokinetics are dose-dependent across the dosing spectrum [10].

This study aimed to formulate an orally fast disintegrating drug delivery system for clonazepam, systematically assessing the influence of various superdisintegrant on disintegration efficiency, dissolution kinetics, and overall bioavailability enhancement to optimize therapeutic efficacy and improve patient adherence.

## 2. Materials

The materials and reagents used in the experiment included: API (Clonazepam; Pharmasia Ltd., Bangladesh), Superdisintegrants (Sodium Starch Glycolate, Kollidone CL, and Ludiflash; Delta Pharmaceutical Ltd., Bangladesh), Diluent (Lactose; Local Market), Binder (Povidone K-30; Local Market), Lubricants (Magnesium Stearate and Talc; Willfrid Smith Ltd, UK), Regents (Sodium Hydroxide and Potassium Dihydrogen Phosphate; Merck, Germany) and Distilled water (Research Laboratory, WUB, Bangladesh).

### 2.1. Equipment’s and Instruments

The equipment and instruments used included: Sieves, Glassware [Morter, Pastel, Beakers, Volumetric flask, Measuring Cylinder, 24 Test Tube, Spatula (large +small), Pipette, Filler] (Local shop), Electronic Balance (Model: Shimadzu, BW 420 H, Japan), Single Punch Compression Machine (Model: Manesty, England), pH meter (Cyber Scan 500, Eutech instruments pte Ltd, Singapore), Digital Slide Calipers (Made in China), Hardness tester (Model: Monsanto hardness tester, India) and Friabilator (Model: Friability test Apparatus (single drum; India), Tablet Dissolution Apparatus (USP-Standard) (Model: Electro lab Tablet Dissolution Tester USPXXIII TDT-06T, India), and UV-Vis-Spectrophotometer (Model: Shimadzu UV Mini 1240, Shimadzu Corporation, Japan).

## 3. Methods

### 3.1. Preformulating Study

Preformulation studies are the first step in dosage form development, focusing on the physicochemical properties of the drug and excipients. These studies guide formulation design, development and manufacturing methods. The following preformulation studies were performed:

#### 3.1.1. Organoleptic properties and Physico-Chemical Evaluations of Clonazepam

Clonazepam was characterized by its organoleptic properties (Clonazepam powder was examined on light and dark backgrounds to observe its physical appearance, color, and odor), melting point (The melting point of clonazepam was determined using the capillary fusion method. A one-sided closed capillary filled with the drug was placed in a melting point apparatus, and the temperature at which the solid drug melted into liquid was recorded), solubility profile (The solubility of clonazepam was assessed by dissolving the drug in various solvents, The solution was filtered, and UV absorbance at 254 nm was recorded after dilution) and moisture content (LOD).

#### 3.1.2. Micrometric properties

According to the reliable reference of the following pre-formulation studies like bulk density, tapped density, compressibility index, Hausner ratio, angle of repose, and porosity were performed for different ingredients of Clonazepam. Tests to evaluate flow ability of a powder [11,12]:

i. **Bulk density:** Defined as mass of many particles of the material divided by total volume (includes particle volume, inter-particle void volume and internal pore volume) they occupy. To determine the bulk density of powder, certain amount of powder should be taken initially then the powder will have to pour into a graduated cylinder and from the cylinder volume have to be estimated.

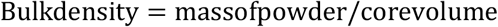
ii. **Tapped density:** Defined as ratio of mass of powder and the tapped volume of the powder. To determine the tapped density of powder, previous amount of powder should be taken in a graduated cylinder and shaken to reduce volume.

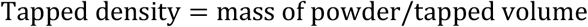
iii. **% Compressibility of powder:** The Carr’s compressibility index is an indication of the compressibility (is the ability of powder to form compactness under pressure) of powder.

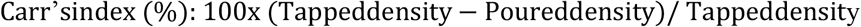
iv. **Hausner ratio:** Value of Hausner ratio less than 1.25 indicates good flow (20% of Carr’s index). Value of Hausner ratio greater than 1.5 indicates poor flow (33% of Carr’s index). Value between 1.25 & 1.5, added glidant to improve flow. Value >1.5, added Glidant does not improve flow.

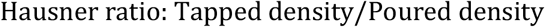
v. **Angle of repose:** Using the funnel, powder is poured slowly & constantly onto the horizontal surface & angle of the resulting pyramid is measured. Mathematically,

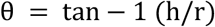

Here, h=height of the pile; r = radius of the pile; and θ = angle of repose.
vi. **Total porosity:** Total porosity [13] was determined by measuring the volume occupied by a selected weight of powder (*V*_*bulk*_) and the true volume of granules (the space occupied by the powder exclusive of spaces greater than the intermolecular space (*V*):

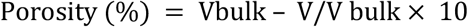

#### 3.1.3. Preparation of Standard Curve of Clonazepam

Standard Clonazepam (working standard) 50 mg was weighed and dissolved in 100 mL of 50 mM phosphate buffer, pH 6.8 to get a concentration of 500 μg/mL of standard stock solution. An ultraviolet spectrophotometric scanning (190-400 nm) was carried out to justify the λ_max_ in the experimental conditions. Then various concentrations ranged between 0-60 μg/mL was prepared from stock solution by serial dilution technique. A calibration curve was obtained by plotting the concentration against absorbance which was taken at λ_max_ (254 nm) using UV spectrophotometer from Shimadzu.

#### 3.1.4. Formulation

In this research work, orally fast disintegrating tablets of clonazepam were designed with an aim of providing faster onset of action by using different superdisintegrants with various ratios (Table: 1).

#### 3.1.5. Preparation of Fast Disintegrating Tablets

Fast disintegrating tablets of clonazepam were prepared by direct compression using varying ratios of superdisintegrants. Ingredients were accurately weighed, sieved for uniform particle size, and mixed for 10 minutes, excluding the lubricant. After adding the lubricant, the mixture was further mixed for 2 minutes and compressed using a hydraulic press with an 8 mm flat punch and dies set.

#### 3.1.6. Physical parameters of resulting tablets

The tablets of each formulation were tested for certain physical parameters e.g., Weight variation, diameter, thickness, friability, potency, hardness,radial tensile strength and axial tensile strength, disintegration time, dispersion time, fitness of dispersion, wetting time, water absorption ratio (R), and *in vitro* dissolution study determination of the prepared tablets etc; in order to ensure that a viable product that is capable of withstanding the rigors of handling and transport is obtained[14].

#### 3.1.7. Dissolution Kinetic modeling

The *in vitro* drug release data of the prepared tablets were analyzed using various kinetic models: Zero order, First order, Higuchi, Korsmeyer Peppas and Hixon-Crowell. Dissolution data were also fitted according to the well-known exponential equation, which is often used to describe the drug release behavior from polymeric systems introduced by Korsmeyer-Peppas.

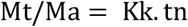

Where, M_t_ is the amount of drug release at time t, M_a_ is the amount of drug release after infinite time; Where K_k_ is the korsmeyer release rate consant and n is the diffusion exponent. Table 2 displayed an analysis of diffusion release mechanism obtained by variyin the n values[15].

**Table 1.** Formulation of clonazepam orally fast disintegrating tablets based on different superdisintegrant.

| Formulation Code | API: Clonazepam | Sodium starch glycolate | Kollidone CL | Ludiflash | Lactose | Povidone K 30 | Magnesium Stearate | Talc | Total weight |
| --- | --- | --- | --- | --- | --- | --- | --- | --- | --- |
| F-1 | 1 | 55 | - | - | 51 | 18 | 10 | 5 | 140 |
| F-2 | 1 | 65 | - | - | 41 | 18 | 10 | 5 | 140 |
| F-3 | 1 | 75 | - | - | 31 | 18 | 10 | 5 | 140 |
| F-4 | 1 | - | 55 | - | 51 | 18 | 10 | 5 | 140 |
| F-5 | 1 | - | 65 | - | 41 | 18 | 10 | 5 | 140 |
| F-6 | 1 | - | 75 | - | 31 | 18 | 10 | 5 | 140 |
| F-7 | 1 | - | - | 55 | 51 | 18 | 10 | 5 | 140 |
| F-8 | 1 | - | - | 65 | 41 | 18 | 10 | 5 | 140 |
| F-9 | 1 | - | - | 75 | 31 | 18 | 10 | 5 | 140 |

*\*Weight of each ingredient was taken in mg*

**Table 2.** Geometric dependence of diffusion expenent n.

| Diffusion Exponent |  |  | Mechanism of transport |
| --- | --- | --- | --- |
| Cylinder | Sphere | Slab |  |
| <0.45 or 0.45 | <0.43 or 0.42 | <0.5 or 0.5 | Fickian (case I) diffusion |
| >0.45 or <0.89 | >0.45 and 0.85 | >0.5 and <1.0 | Anomalous/Non-Fickian transport |
| 0.89 | 0.85 | 1.0 | Case II/Zero order transport |
| >0.89 | >0.85 | >1.0 | Super Case II transport |

#### 3.1.8. Dissolution efficiency (DE)

The drug-release profiles were characterized by calculating the DE, which is defined as the area under the dissolution curve up to a certain time t_1_, expressed as a percentage of the area of the rectangle arising from 100% dissolution at the start, extended over the same time period; where y is the percent drug dissolved at time t_1_[16].DE can be calculated as a fraction (from 0 to 1) by the following equation:

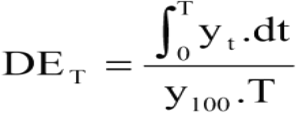

#### 3.1.9. Similarity factor (f_2_)

The comparison of dissolution profiles between marketed product and formulations were evaluated. To statistically assess the similarity between these dissolution profiles, the similarity factor (f_2_) was calculated [17].

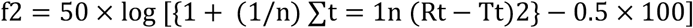

Where, n= number of time points; R_t_= cumulative percentage dissolved of the reference (marketed) product at time t and T_t_= cumulative percentage dissolved of the test (faster release formulation of glimepiride) product at time t.

The f_2_ value of 100% indicates identical dissolution profiles between the test and reference. 50 < f_2_ < 100 suggests similarity between the profiles, while a smaller f_2_ indicates greater dissimilarity in release rates [18].

#### 3.1.10. Stability studies

The FDA and ICH provide guidelines for stability testing of new drug products, which are required for pharmaceutical registration for the purpose of human use. According to ICH, long-term stability testing should be conducted at 25°C / 60% RH for 12 months, while accelerated testing should be at 40°C / 75% RH for six months. Intermediate stability testing should be at 30°C / 65%RH. Various storage conditions and testing periods are outlined in Table 3.

**Table 3.** ICH guideline for stability study.

| Study | Storage condition | Duration |
| --- | --- | --- |
| Long-term | 25°C ± 2°C, 60% ± 5% RH | 12 months |
| Intermediate | 30°C ± 2°C, 65% ± 5% RH | 6 months |
| Accelerated temperature | 40°C ± 2°C, 75% ± 5% RH | 6 months |

## 4. Results and Discussion

### 4.1. Standard Curve of Clonazepam

A calibration curve for clonazepam (*y* = 0.0471*x*+ 0.0127, *R*2 = 0.997) was obtained by plotting the concentration against absorbance which was taken at λ_max_ (254 nm) using UV spectrophotometer from Shimadzu (Figure 1).

**Fig 1.**
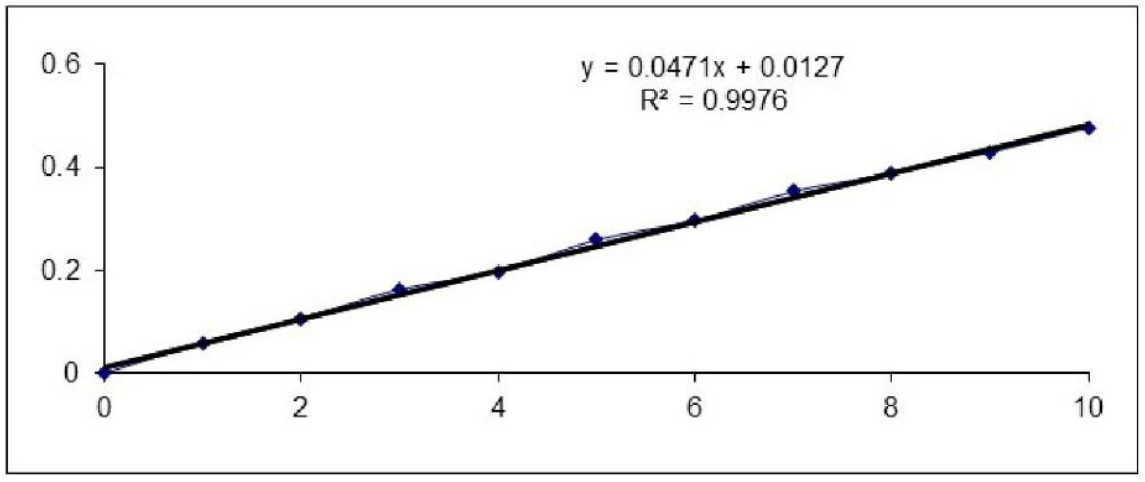
Standard calibration curve of clonazepam recorded by UV spectrophotometer at 254 nm.

### 4.2. Characterization of drug

Organoleptic properties like (appearance, color, and odor) and various physico-chemical evaluations of clonazepam were represented in Table 4.

**Table 4.** Organoleptic and physico-chemical evaluations of clonazepam.

| Parameters | Interpretation |
| --- | --- |
| Appearance | Crystalline powder |
| Color | Slightly yellowish |
| Odor | Odorless |
| Solubility | Practically insoluble in water, freely soluble in methanol |
| Melting point | 239° C |
| LOD | Maximum 0.42 % |

### 4.3. Evaluation of powder blends

Flow properties of different formulations are showed in (Table 5). Carr’s index and Hausner’s ratio were calculated. The percent compressibility lies within the range of (14.21-20.55) % and Hausner’s ratio is in the range of (1.12-1.19) for different ingredients respectively. Different formulations showed the angle of repose within 25^0^. The percentage porosity values of the granules ranged from (12.2% to 17.4%) indicating that the packing of the granules may range from close to loose packing and also further confirming that the particles are not of greatly different sizes. The overall results confirm that the powder blends demonstrate appropriate flow and packing characteristics, essential for achieving uniform die filling, minimizing weight variation, and ensuring reproducible tablet properties [19]. These findings validate the formulation approach, highlighting its potential for optimizing clonazepam OFDT production with efficient manufacturability and performance.

**Table 5.** Evaluation of powder flow properties of different formulations of clonazepam.

| Formulation Code | Carr's index (%) | Hausner's ratio | Angle of repose ($^{\circ}$C) | Porosity % |
| --- | --- | --- | --- | --- |
| F-1 | 14.79 | 1.16 | 21 | 14.8 |
| F-2 | 17..35 | 1.18 | 20 | 16.1 |
| F-3 | 19.11 | 1.19 | 24 | 13.9 |
| F-4 | 15.88 | 1.12 | 23 | 12.2 |
| F-5 | 16.72 | 1.13 | 20 | 16.7 |
| F-6 | 18.41 | 1.17 | 22 | 15.3 |
| F-7 | 14.21 | 1.13 | 21 | 12.5 |
| F-8 | 18.63 | 1.15 | 23 | 17.4 |
| F-9 | 20.55 | 1.14 | 24 | 15.6 |

### 4.4. Physical parameters of Clonazepam Orally Fast Disintegrating Tablets

The average weight variation, diameter, thickness, friability and potency (calculated by comparing the actual drug content with the expected content, and expressed as a percentage) of the prepared tablets from formulations F-1 to F-9 are presented in Table 6.

**Table 6.**
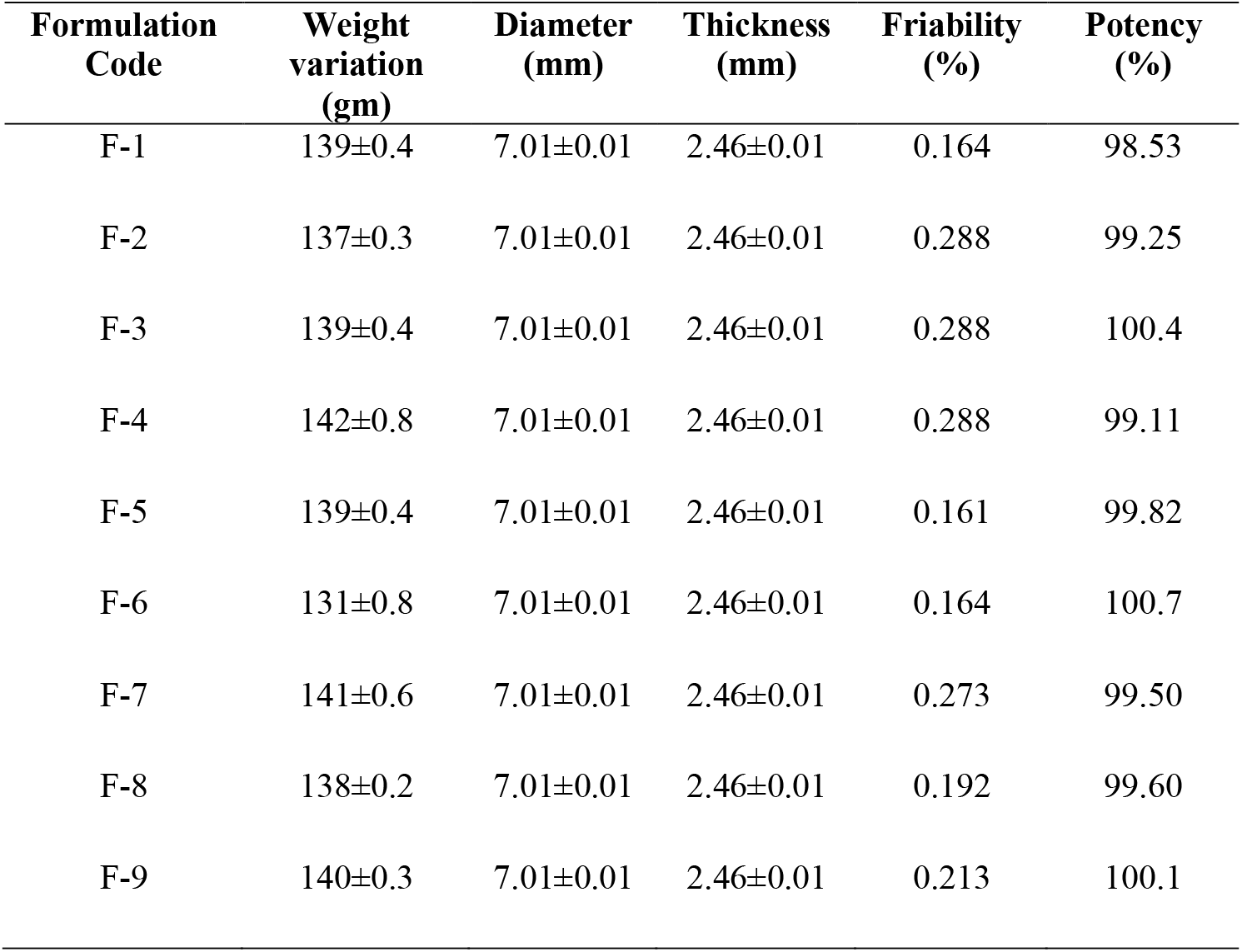
Average weight variation, diameter, thickness, and friability of the prepared tablets from formulations F-1 to F-9 (Number of samples=20)

| Formulation Code | Weight variation (gm) | Diameter (mm) | Thickness (mm) | Friability (%) | Potency (%) |
| --- | --- | --- | --- | --- | --- |
| F-1 | 139±0.4 | 7.01±0.01 | 2.46±0.01 | 0.164 | 98.53 |
| F-2 | 137±0.3 | 7.01±0.01 | 2.46±0.01 | 0.288 | 99.25 |
| F-3 | 139±0.4 | 7.01±0.01 | 2.46±0.01 | 0.288 | 100.4 |
| F-4 | 142±0.8 | 7.01±0.01 | 2.46±0.01 | 0.288 | 99.11 |
| F-5 | 139±0.4 | 7.01±0.01 | 2.46±0.01 | 0.161 | 99.82 |
| F-6 | 131±0.8 | 7.01±0.01 | 2.46±0.01 | 0.164 | 100.7 |
| F-7 | 141±0.6 | 7.01±0.01 | 2.46±0.01 | 0.273 | 99.50 |
| F-8 | 138±0.2 | 7.01±0.01 | 2.46±0.01 | 0.192 | 99.60 |
| F-9 | 140±0.3 | 7.01±0.01 | 2.46±0.01 | 0.213 | 100.1 |

The average tablet weight for all formulations was consistently close to the theoretical value of 140 mg. The average Diameter was also found to be much consistent varying between the ranges of 7.01mm. The average Thickness of the tablets also ranged between 2.46mm; however, the variations were not alarming and remained within the acceptable range. The friability of the tablets varied significantly, ranging from 0.161% to 0.288%, with the highest friability observed in formulations F2, F3, and F4, indicating maximum loss upon attrition. According to references [20], the acceptable friability range is 0.5-1%. Since no formulation exceeded 1%, friability is not a major concern. The potency test results ranged from 98.53% to 100.7%, with Formulations F-3, F-6,andF-9 showing the highest potency, while F-1 had the lowest. All formulations complied with the B.P. specifications for potency, indicating that they met the required standards for drug content.

Hardness of the tablet is also expressed in two terms: radial tensile strength and axial tensile strength. Radial tensile strength is the force required to break a tablet. The Radial tensile strength, T of the tablets can be calculated from the equation:

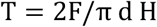

Where, F is the load needed to break the tablet and d and H are the diameter and thickness respectively.

The Axial tensile strength (T_x_) can be calculated from the following relationship:

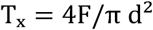

Hardness, diameter and thickness of the tablets are factors of radial tensile strength and hardness and diameter measures for the axial tensile strength.

Hardness, radial tensile strength and axial tensile strength of the tablets of different formulations F-1 to F-9 are presented in Table 7.

**Table 7.**
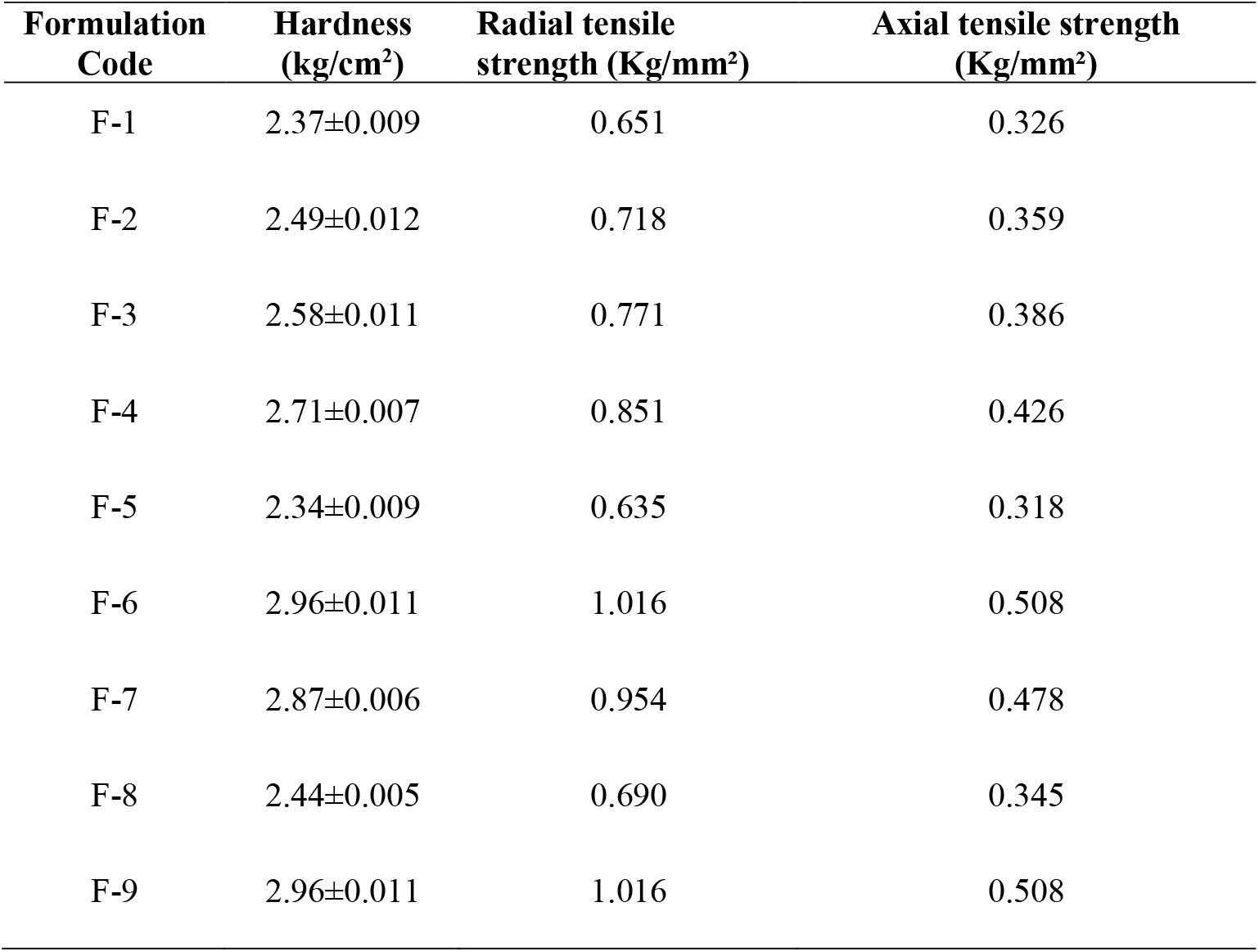
Average weight variation, diameter, thickness, and friability of the prepared tablets from formulations F-1 to F-9 (Number of samples=20)

The hardness values of the tablet formulations ranged from (2.34 to 2.96 kg/cm^2^), indicating uniformity and consistency. The radial tensile strength (RTS)and axial tensile strength (ATS) values for all formulations fell within acceptable ranges, with RTS values ranging from (0.635 to 1.016 Kg/mm^2^) and ATS values ranging from (0.318 to 0.508 Kg/mm^2^). These results suggested that the tablets have good mechanical strength, with none of the formulations showing values outside typical limits, ensuring their suitability for further development, use and highlights their superior mechanical properties, which can be attributed to the optimized excipient selection and formulation techniques[21].The findings indicate that the balance between mechanical strength and rapid disintegration was effectively maintained, making these formulations viable applicants for further optimization and therapeutic application[22].

Disintegration Time, Dispersion Time, Fitness of dispersion, Wetting time and Water absorption ratio of the prepared tablets of different formulations (F-1 to F-9) are presented in Table 8.

**Table 8.** Disintegration Time, Dispersion Time, Fitness of dispersion, Wetting time and Water absorption ratio of the prepared tablets of different formulations (F-1 to F-9)

| Formulation Code | Disintegration time (sec) | Dispersion time (sec) | Fitness of dispersion | Wetting time (sec) | Water absorption ratio |
| --- | --- | --- | --- | --- | --- |
| F-1 | 33 | 195 | Passed | 32 | 61.33 |
| F-2 | 27 | 180 | Passed | 26 | 42.96 |
| F-3 | 20 | 140 | Passed | 22 | 21.45 |
| F-4 | 42 | 230 | Passed | 57 | 77.96 |
| F-5 | 30 | 210 | Failed | 49 | 54.32 |
| F-6 | 22 | 190 | Passed | 35 | 27.69 |
| F-7 | 47 | 167 | Failed | 54 | 71.32 |
| F-8 | 28 | 214 | Passed | 26 | 50.68 |
| F-9 | 33 | 159 | Passed | 31 | 29.12 |

**Table 9.** Release rate constants and Correlation coefficient (R^2^) values of different formulations (F-1 to F-9)

| Formulation Code | Zero Order |  | First Order |  | Higuchi |  | Korsmeyer-Peppas |  | Hixson-Crowell |  |
| --- | --- | --- | --- | --- | --- | --- | --- | --- | --- | --- |
| | $K_0$ | $R^2$ | $K_1$ | $R^2$ | $K_H$ | $R^2$ | n | $R^2$ | $K_{H-C}$ | $R^2$ |
| F-1 | 84.51 | 0.873 | -1.087 | 0.987 | 96.31 | 0.984 | 0.080 | 0.043 | 3.103 | 0.541 |
| F-2 | 82.41 | 0.799 | -1.402 | 0.982 | 97.24 | 0.966 | 0.064 | 0.051 | 2.846 | 0.444 |
| F-3 | 77.41 | 0.702 | -1.677 | 0.958 | 94.89 | 0.916 | 0.052 | 0.055 | 2.793 | 0.406 |
| F-4 | 84.79 | 0.881 | -1.003 | 0.988 | 95.07 | 0.984 | 0.078 | 0.039 | 3.109 | 0.550 |
| F-5 | 81.37 | 0.814 | -1.180 | 0.979 | 95.28 | 0.969 | 0.064 | 0.042 | 2.959 | 0.481 |
| F-6 | 75.94 | 0.714 | -1.251 | 0.968 | 92.65 | 0.922 | 0.048 | 0.042 | 2.787 | 0.413 |
| F-7 | 87.10 | 0.882 | -1.200 | 0.992 | 98.91 | 0.987 | 0.084 | 0.048 | 3.139 | 0.547 |
| F-8 | 83.22 | 0.803 | -1.146 | 0.966 | 98.67 | 0.972 | 0.074 | 0.053 | 3.023 | 0.494 |
| F-9 | 75.94 | 0.714 | -1.251 | 0.968 | 98.00 | 0.966 | 0.068 | 0.054 | 2.966 | 0.472 |

Disintegration time is very important for tablets dosage form as the internal structure of tablets (pore size distribution, water penetration capability and swelling of disintegrating substance) suggested the mechanism of disintegration. Disintegration time of formulated tablets was within the range. All the formulations show disintegration time less than 60 seconds whereas kollidon CL and ludiflash showed more disintegration time than sodium starch glycolate. The dispersion time was also found to be varying between the ranges of (140-230) seconds. Fitness of dispersion test of the tablets passed for all formulations; excluding both the formulations (F-5, and F-7). Wetting time of different formulations ranged from (22-57) seconds and also water absorption ratio for different formulations were estimated from (21.45 to 77.96). Formulations with higher water absorption ratios tend to have slower disintegration and dispersion times, as excessive water uptake can lead to gelation or excessive swelling that slows down disintegration rather than facilitating it [23]. Though the variations of abovementioned tests were varied within the acceptable range.

### 4.5. Dissolution Studies of the tablets of different formulations

Clonazepam orally fast disintegrating tablets were subjected to various kinetic models, including Zero order, First order, Higuchi, Korsmeyer-Peppas, and Hixson-Crowell (Figures 2 to 4).

**Fig 2.**
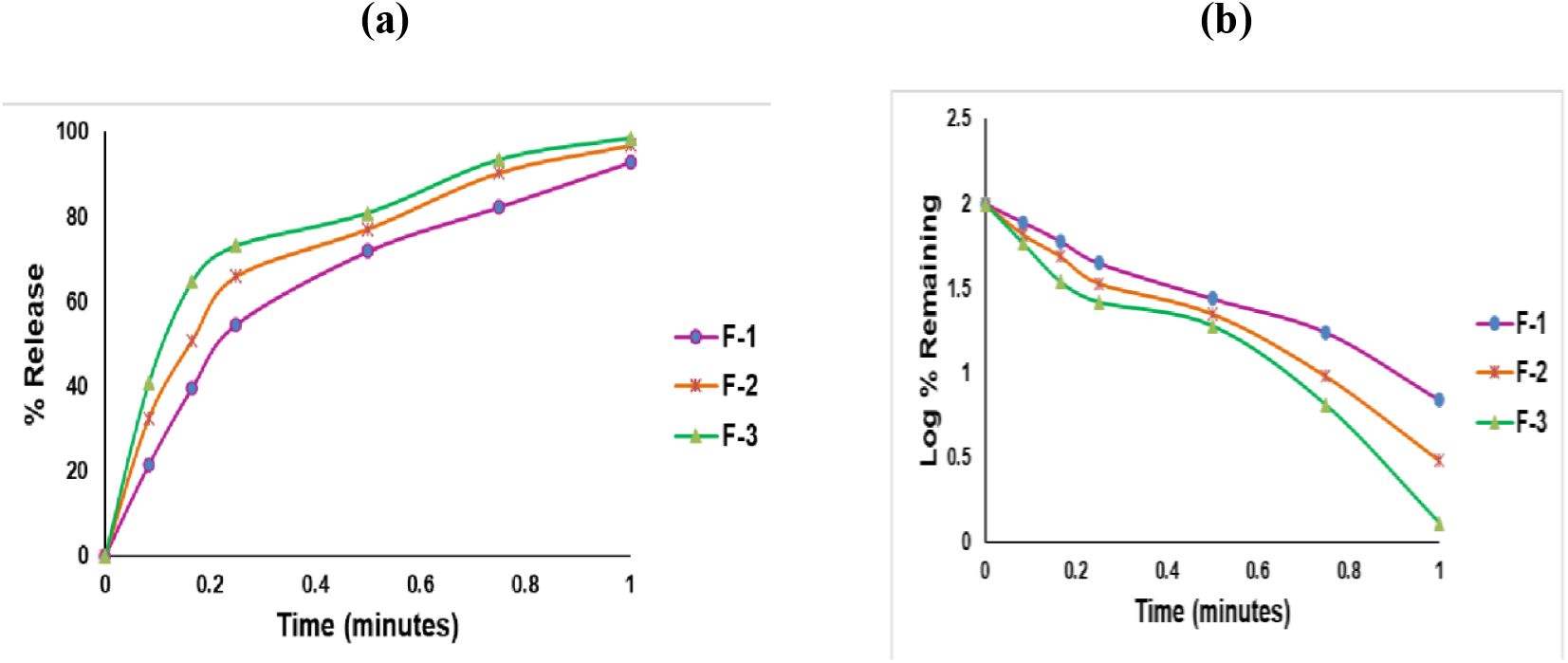
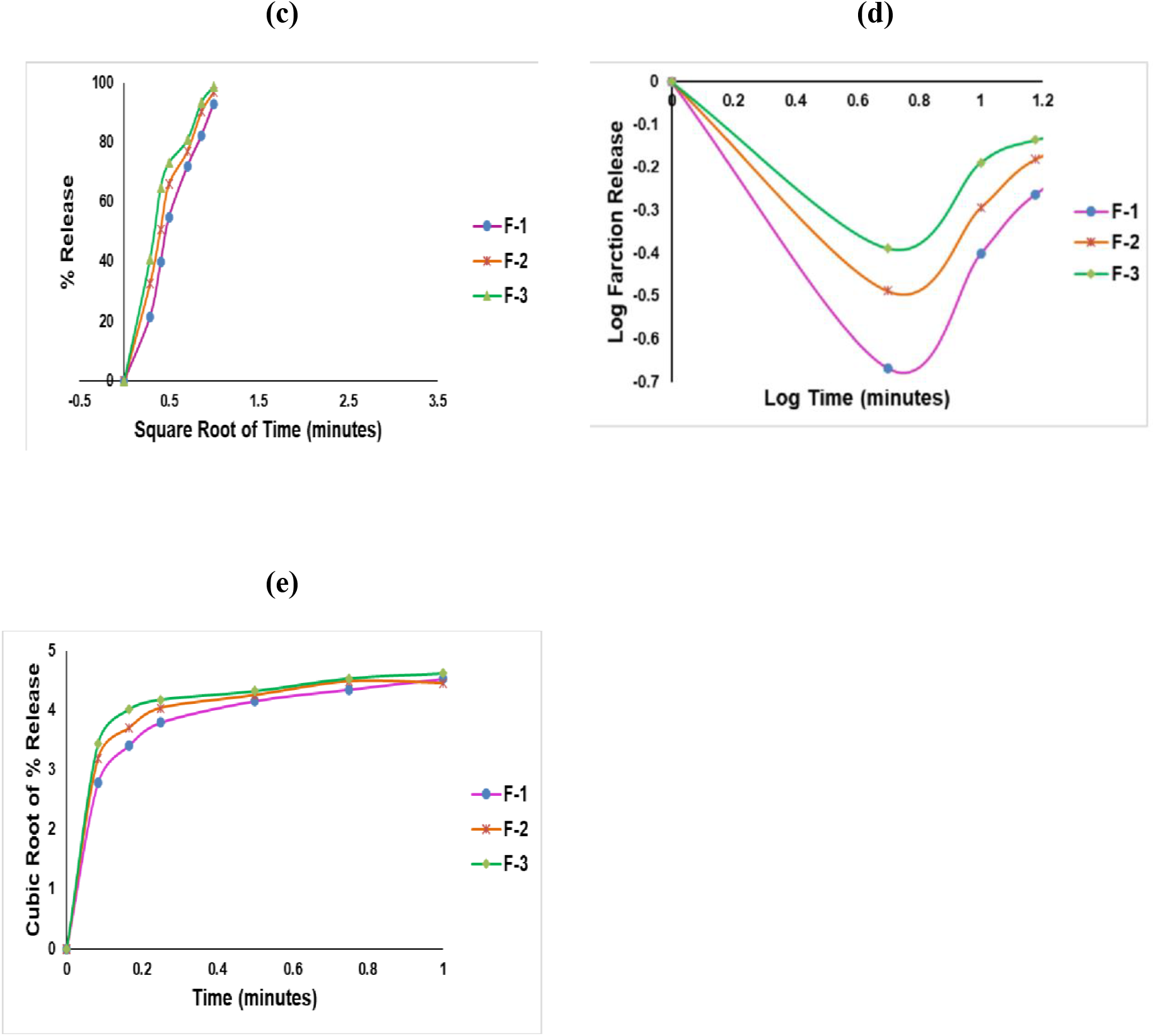
Release kinetics of clonazepam orally fast disintegrating tablets of different formulations (F-1, F-2, and F-3): a) Zero order, b) First order, c) Higuchi model, d) Korsmeyer-Peppas model, and e) Hixson-Crowell model

**Fig 3.**
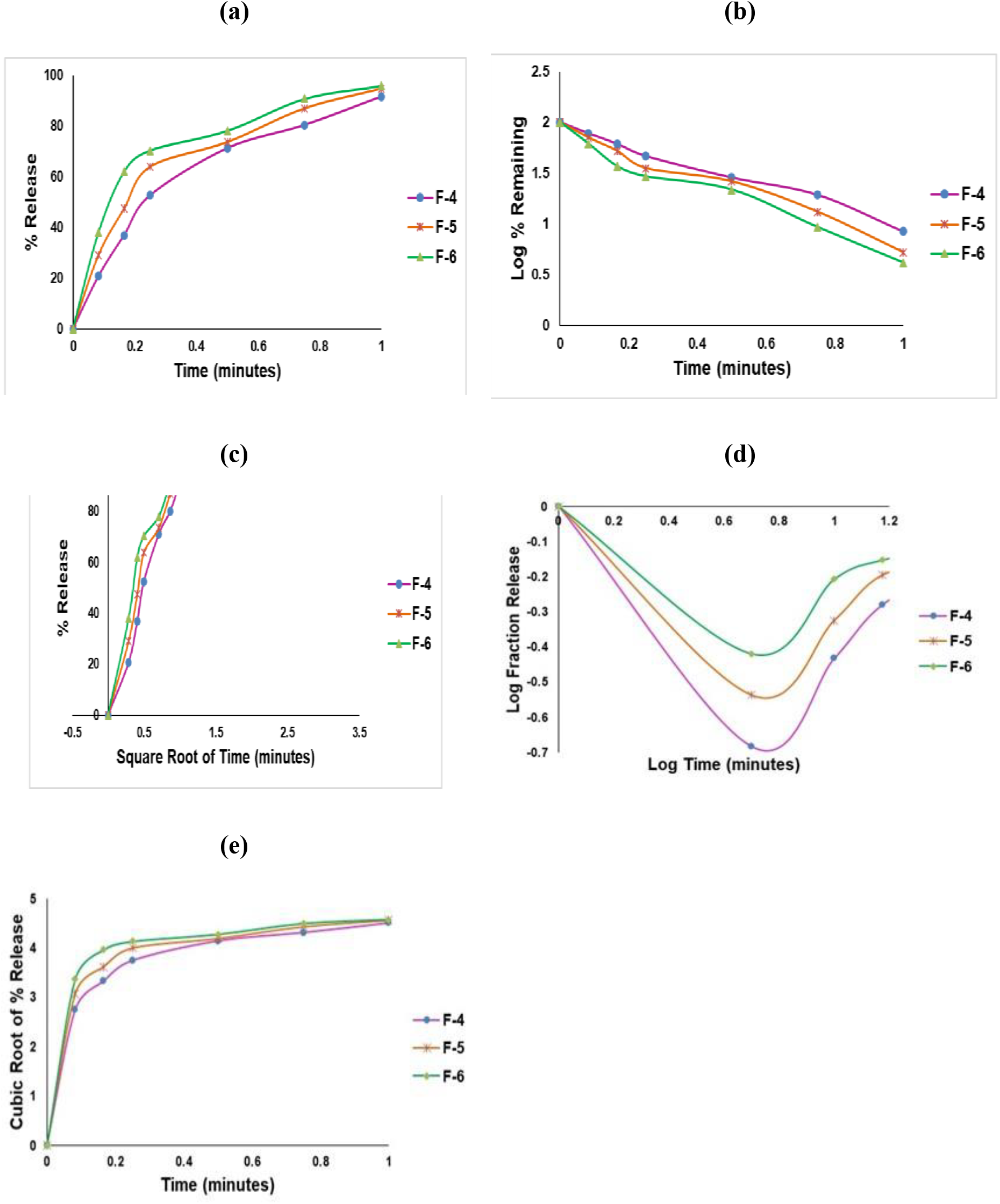
Release kinetics of clonazepam orally fast disintegrating tablets of different formulations (F-4, F-5, and F-6): a) Zero order, b) First order, c) Higuchi model, d) Korsmeyer-Peppas model, and e) Hixson-Crowell model

**Fig 4.**
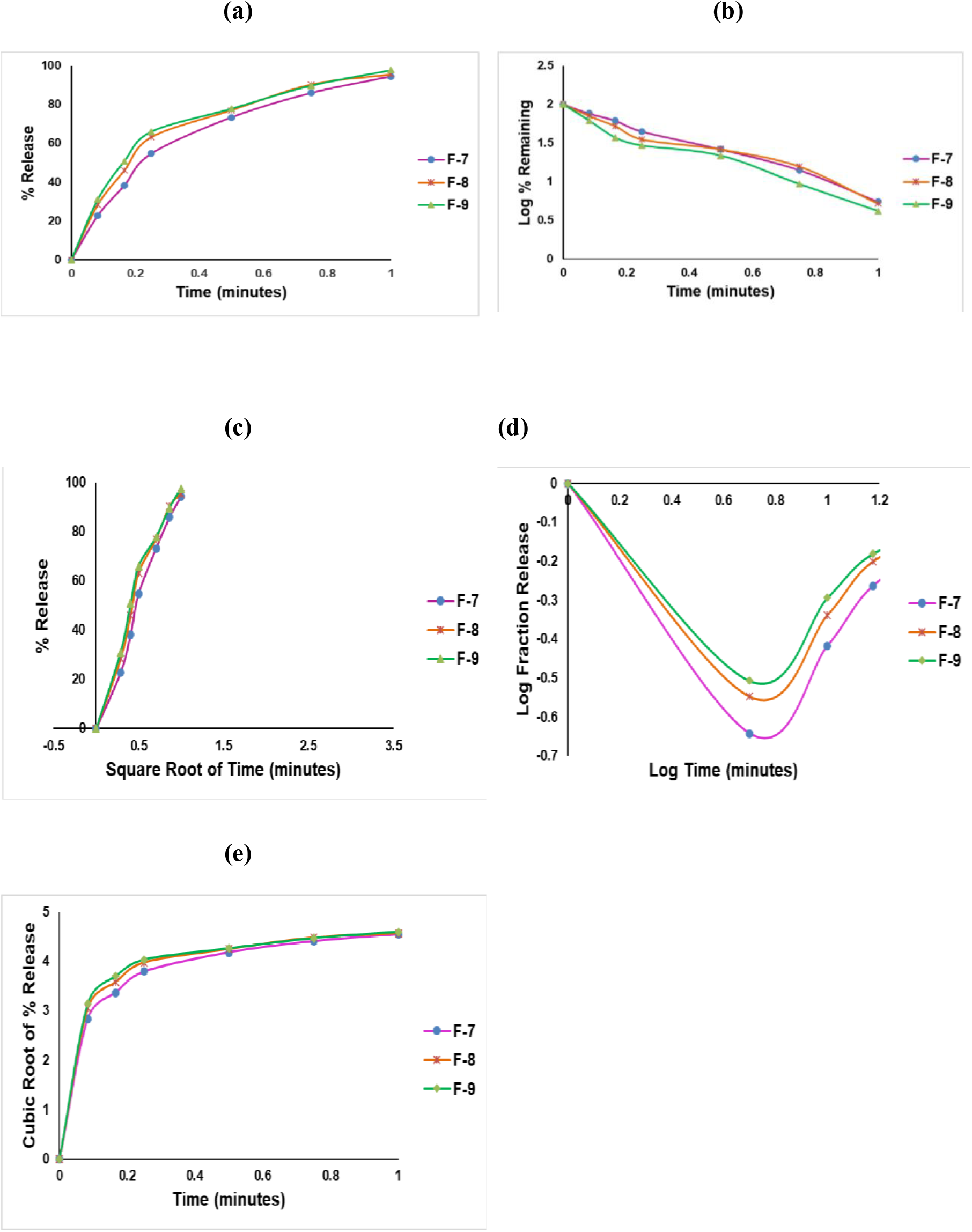
Release kinetics of clonazepam orally fast disintegrating tablets of different formulations (F-7, F-8, and F-9): a) Zero order, b) First order, c) Higuchi model, d) Korsmeyer-Peppas model, and e) Hixson-Crowell model

The study assessed drug release kinetics using various plots such as zero-order kinetics (percent release vs. time)[24], showed F-3 released 98.7% at 60 minutes, while F-4 released 91.47%; first-order kinetics (log percent drug remaining vs. time) revealed F-4 had a log % remaining of 0.93, and F-3 had 0.11 at 60 minutes; Higuchi plot (percent release vs. square root of time) indicated F-3 released 98.7% at 60 minutes, while F-4 released 91.47%; Korsmeyer release profile(log fraction release vs. log time) showed F-3 had a log fraction release of −0.00568, and F-4 had −0.03868 60 minutes; and Hixson-Crowell profile (cubic root of % release vs. time) displayed F-4 with a cubic root value of 4.6213, compared to F-3 4.5057. These values indicate that formulation F-3 demonstrated faster and higher drug release than formulation F-4.

Clonazepam orally fast disintegrating tablets release profiles were analyzed using various kinetic models. Formulation F-7 demonstrated the most favorable results, showing a first order release kinetics with a high R^2^ value of 0.992, indicating concentration-dependent drug release. The release mechanism followed anomalous (non-Fickian) diffusion, as suggested by the Korsmeyer-Peppas model with an “n” value of 0.084. This indicates that both diffusion and erosion contribute to the drug release process. Overall, F-7 exhibited optimal drug release behavior, making it the most promising formulation among the tested samples; suggests that drug release is governed by both diffusion and polymer erosion [25].

### 4.6. Dissolution efficiency (DE) of the formulations

The Dissolution Efficiency (DE) of nine formulations (F-1 to F-9) was evaluated. F-5 showed the highest DE at 94%, indicating superior dissolution. F-6 (88%) and F-8 (87%) also exhibited good dissolution profiles. F-7, F-9, andF-3 had moderate DE values (85%-84%), while F-1, F-2 and F-4 displayed the lowest efficiencies (80% and 83%, respectively). The results suggest that F-5 is the most efficient formulation, while F-1 requires optimization. These findings highlight the importance of formulation adjustments to improve dissolution and bioavailability of a drug, as a higher DE ensures better solubility and absorption in the gastrointestinal tract [26].

#### Similarity factor (f_2_)

The similarity factor (f_2_) was calculated to compare the drug release profiles of nine proposed formulations with the marketed product. Formulations (F-1, F-3, F-6, F-8, andF-9) exhibited moderate similarity with (f_2_)values ranging from 20 to 30, indicating some resemblance to the marketed profile but still showing room for improvement. In contrast, formulations (F-2, F-4, F-5, and F-7) displayed low similarity with (f_2_) values below 20, suggesting significant deviations in release behavior.

### 4.7. Stability testing of the best formulation

Under different storage conditions and durations of the stability study, all parameters, including drug content and dissolution profiles, remained consistent with initial values. No significant changes were observed in any of the tested values or the physical appearance of the tablets. The stability of clonazepam FDTs prepared with super disintegrants holds significant importance in the pharmaceutical field [27]. The use of super disintegrants enhances the rapid breakdown and dissolution of the tablet in the mouth, leading to quicker absorption of clonazepam in the bloodstream [28]. This is particularly beneficial for patients suffering from anxiety or seizures who require immediate relief [29]. Additionally, the stability of the formulation ensures that the active ingredient and the super disintegrants remain effective and do not degrade over time [30].

Therefore, it can be concluded that the clonazepam oral fast-dissolving tablets prepared with super disintegrants were stable.

## 5. Conclusion

In conclusion, the study demonstrated that all clonazepam tablet formulations, incorporating various excipients, met the required characterization and evaluation standards. Formulations containing superdisintegrants, particularly F-3, F-6, and F-9, showed superior release profiles with faster drug release, which was attributed to the specific characteristics and concentrations of the superdisintegrants used. Overall, formulation F-5 demonstrated the highest dissolution efficiency, while formulations like F-1, F-2, and F-4 require optimization. The similarity factor analysis showed moderate alignment with the marketed product for some formulations, indicating potential for further refinement. Stability testing confirmed the consistency and safety of all formulations, with no significant changes observed. These findings underscore the significance of strategic superdisintegrants selection in optimizing OFDT formulations for improved therapeutic efficacy and patient adherence. Further research may explore refining formulation parameters to enhance bioavailability and commercial viability.

## Data Availability

All data are available in the manuscript.

## Acknowledgment

The authors gratefully acknowledge the support provided by Pharmasia Ltd., Bangladesh and Delta Pharmaceutical Ltd., Bangladesh, and the use of the Research Pharmaceutical Lab, Department of Pharmacy, World University of Bangladesh, for their invaluable contributions to this study.

## Author contribution

All authors have equal contribution

